# COVID-19: Impact of Obesity and Diabetes in Disease Severity

**DOI:** 10.1101/2020.05.24.20111724

**Authors:** Salman Al-Sabah, Mohannad Al-Haddad, Sarah Al-Youha, Mohammad Jamal, Sulaiman Almazeedi

**Affiliations:** COVID-19 Research Group, Jaber Al-Ahmad Al-Sabah Hospital, Kuwait; Kuwait University, college of Medicine

## Abstract

**Background:** The Coronavirus disease 2019 (COVID-19) pandemic is straining the healthcare system, particularly for patients with severe outcomes who require admittance to the intensive care unit (ICU). This study aimed to investigate the potential associations of obesity and diabetes with COVID-19 severe outcomes, assessed as ICU admittance.

**Subjects:** Demographic and patient characteristics from a retrospective cohort of 1158 patients hospitalized with COVID-19 in a single center in Kuwait, along with their medical history, were analyzed. Univariate and multivariate analyses were performed to explore the associations between different variables and ICU admittance.

**Results:** From the 1158 hospitalized patients, 271 (23.4%) had diabetes, 236 (20.4%) had hypertension and 104 (9%) required admittance into the ICU. From patients with available measurements, 157 (21.6%) had body mass index (BMI)≥25 kg/m^2^. Univariate analysis showed that overweight (BMI=25.0–29.9 kg/m^2^), obesity class I (BMI=30–34.9 kg/m^2^) and morbid obesity (BMI≥40 kg/m^2^) associated with ICU admittance (odds ratio (OR) [95% confidence intervals (CI)]: 2.45 [1.26–4.74] *p*-value=0.008; OR [95% CI]: 3.51 [1.60–7.69] *p*-value=0.002; and OR [95% CI]: 5.18 [1.50–17.85] *p*-value=0.009], respectively). Patients with diabetes were more likely to be admitted to ICU (OR [95% CI]: 9.38 [5.49–16.02]). Two models for multivariate regression analysis were used, assessing either BMI or diabetes on ICU outcomes. In the BMI model, class I obesity and morbid obesity were associated with ICU admittance (adjusted OR (AOR) [95% CI]: 2.7 [1.17–6.20] *p*-value=0.019 and AOR [95% CI]: 3.95 [1.00–15.20] *p*-value=0.046, respectively). In the diabetes model, diabetes was associated with higher ICU admittance (AOR [95% CI]: 5.49 [3.13–9.65] *p*-value<0.001) whereas hypertension had a protective effect on ICU admittance (AOR [95% CI]: 0.51 (0.28–0.91).

**Conclusions:** In our cohort, overweight, obesity and diabetes in patients with COVID-19 were associated with ICU admittance, putting these patients at higher risk of poor outcomes.

## Introduction

The outbreak and unprecedented spread of SARS-CoV-2, responsible for COVID-19, has taken the entire world by storm since December 2019. On March 11, 2020, the World Health Organisation labelled the COVID-19 outbreak a pandemic(1).

The novel coronavirus SARS-CoV-2 appears to affect some people more severely than others. Most patients with SARS-CoV-2 infection have only mild or no symptoms, thus considered to have a mild form of the disease. However, some patients have developed life-threatening acute respiratory distress syndrome, septic shock and multiorgan failure, including acute renal failure and cardiac injury caused by a cytokine storm, which increases the risk of mortality by 15% to 20% (2).

Various risk factors have been linked to the progression of COVID-19. Recognition of such factors can help to highlight a high-risk population and determine prevention strategies. Old age, chronic disease, respiratory disease and cardiovascular disease have been studied intensively and found to have a significant association with the severity of COVID-19 (3). Patients with obesity and diabetes are also at risk of more severe outcomes of COVID-19, which is particularly important to healthcare workers considering the high prevalence of these conditions in the Middle East, Europe, and the United States.

In this study, we explored the potential association of obesity and diabetes with severe outcomes in patients hospitalized with SARS-CoV-2 infection in Kuwait.

## Subjects and Methods

### Study design and data collection

Ethical approval for the study was obtained from the Ministry of Health in Kuwait. This retrospective cohort study included 1158 patients previously diagnosed with COVID-19, who were admitted to the Jaber Al-Ahmad Al-Sabah Hospital in Kuwait from 24 February to 7 April 2020. Jaber Hospital is a 1168 bed hospital that has become entirely dedicated for COVID-19 patients. It is unique in that it has been used as both a treatment site and an institutional quarantine facility by the Kuwaiti government. Prior to the first case of COVID-19 even been diagnosed in Kuwait, on February 24^th^, 2020, all incoming travellers to Kuwait were screened and admitted to Jaber Al-Ahmad Al-Sabah hospital irrespective of symptomatic status. As a result, the COVID-19 patient population at Jaber Hospital is composed of a heterogenous patient population in terms of clinical presentation and demographic profile. Testing for COVID-19 was conducted via real-time reverse-transcription polymerase chain reaction assays of nasal swab specimens. Only patients with positive results were included in the study; patients with negative or equivocal results were excluded.

Demographic data, including age, sex, body mass index (BMI), fasting blood glucose level, systolic blood pressure, diastolic blood pressure and past medical history, were extracted from the Jaber Al-Ahmed Al-Sabah hospital’s electronic medical records. To maintain confidentiality, these data were de-identified.

Subjects were classified based on their body mass index (BMI), as follows: normal weight (BMI of 18.5–24.9 kg/m^2^), overweight (BMI of 25.0–29.9 kg/m^2^) and obese (BMI ≥30 kg/m^2^. Obese subjects were further stratified into classes: class I obesity was defined as a BMI of 30–34.9 kg/m^2^; class II obesity, by a BMI of 35–39.9 kg/m^2^; and morbid obesity, by a BMI ≥40 kg/m^2^. Hypertension was defined as systolic blood pressure of ≥130 mm Hg or diastolic blood pressure of ≥85 mm Hg. Prediabetes was defined as a fasting blood glucose level of 100–126 mg/dL, and diabetes was defined as a fasting blood glucose level of ≥126 mg/dL.

COVID-19 progression was defined as a patient’s admission to the intensive care unit (ICU) for more active systemic treatment, such as systemic glucocorticoids or intravenous immunoglobulin, or for further advanced respiratory support, such as mechanical ventilation or extracorporeal membrane oxygenation.

### Statistical analysis

Categorical variables were summarised as percentages and analysed with the chi-square test. Continuous variables were summarised as medians with interquartile ranges (IQRs) and compared in Student’s *t* test or the nonparametric Mann-Whitney *U* test. Each promising variable with a plausible biological reason for inclusion in further analysis was verified for normal distribution using the Shapiro-Wilk and Kolmogorov-Smirnov tests. Continuous score variables were set to 0 or 1 to represent the values below or above a predefined threshold based on the literature; binary variables were also set at 0 or 1 (absent versus present, respectively).

Potential predictors for admission into the ICU were assessed using univariate and multivariate logistic regression analyses, and logistic regression models were adjusted by age and sex. Considering the association between obesity and diabetes, we constructed two multivariate logistic regression models to predict the independent associations of each more accurately of these two conditions with severe outcomes of COVID-19 (ICU admittance). The associations between the exposure and outcomes were expressed in terms of the odds ratio (OR), along with 95% confidence intervals (Cis). Goodness-of-fit analyses of the models were done using the C-statistic (area under the receiver operating characteristic [ROC] curve) and 95% CI. A *p* value of 0.05 was considered statistically significant for all the statistical tests. All statistical analyses were performed with R software (R Project for Statistical Computing, Vienna, Austria; R Core Team, 2019).

## Results

### Characteristics of hospitalized patients

From 24 February to 5 May 2020, 1158 consecutive patients with COVID-19 were treated at Jaber Al-Ahmed Al-Sabah Hospital. Of these, 945 (81.6%) were male (mean age ± standard deviation [SD]: 41.5 ± 13.5 years), and 213 (18.4%) were female (mean age ± SD: 44.8 ± 18.9 years). The majority of patients were either of Indian nationality (n=550, 47.5%) or Kuwaiti nationals (n=301, (26%). The most common symptoms of COVID-19 were cough (n = 344, 29.7%), chills (n = 327, 28.2%) and sore throat (n = 135, 11.7%).

The baseline characteristics of the study patients are listed in **Table 1**. From patients with BMI measurements taken at baseline (n=727), 461 patients (39.8%) had BMI ≥ 25 kg/m^2^: 304 patients were overweight, 89 had class I obesity, 40 had class II obesity and 19 had morbid obesity. Blood glucose measurements revealed that 314 (27.1%) of the participants had prediabetes and 271 (23.4%) had diabetes. Hypertension was diagnosed in 236 patients (20.4%). By the time of the analysis, 104 patients (9%) had been admitted to the ICU for treatment of COVID-19, and there had been a total of 40 deaths (**Table 1**).

**Table 1:**
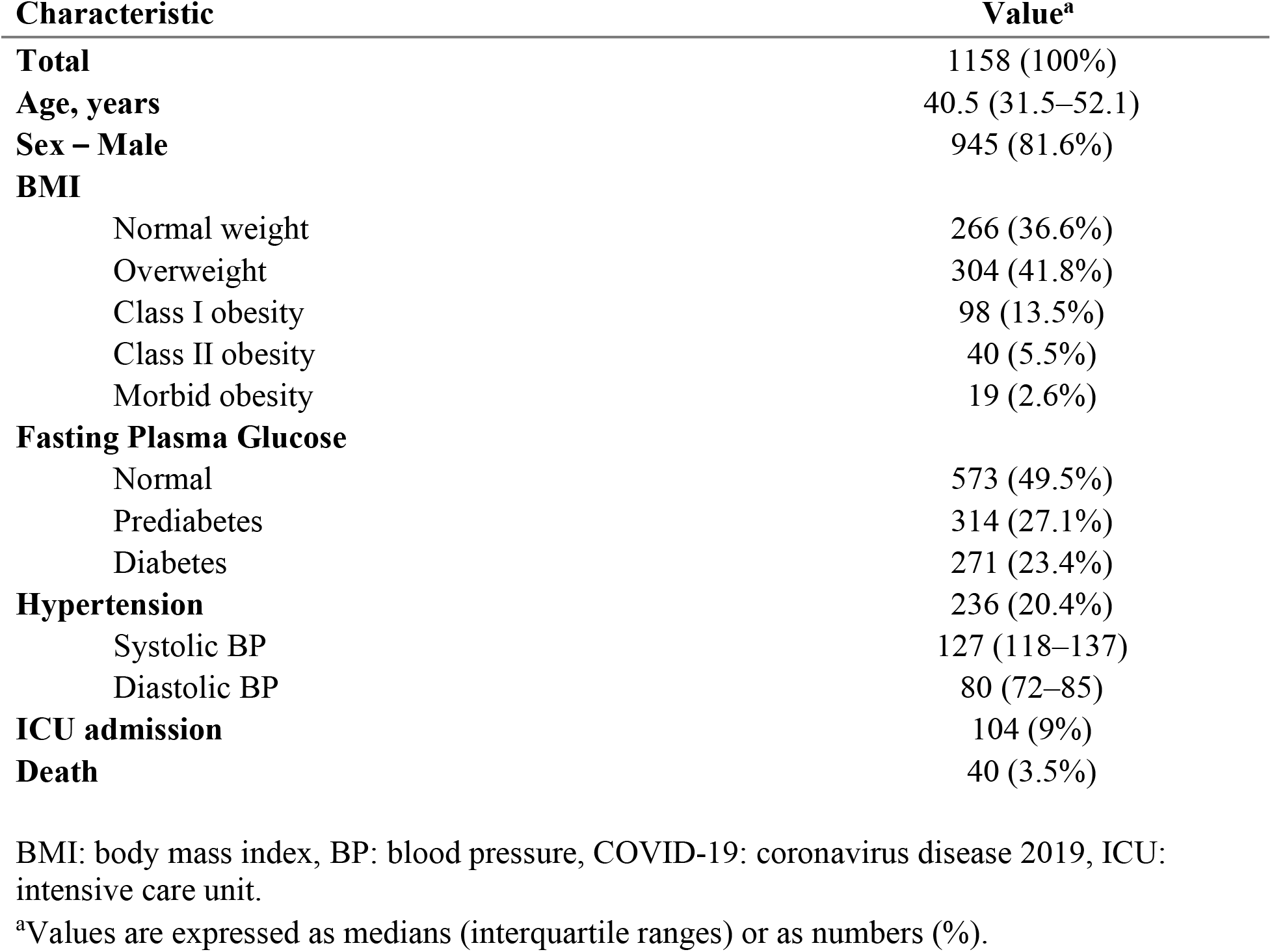
Baseline characteristics of patients hospitalized with COVID-19.

| **Characteristic** | **Value<sup>a</sup>** |
| --- | --- |
| **Total** | 1158 (100%) |
| **Age, years** | 40.5 (31.5–52.1) |
| **Sex – Male** | 945 (81.6%) |
| **BMI** |  |
| Normal weight | 266 (36.6%) |
| Overweight | 304 (41.8%) |
| Class I obesity | 98 (13.5%) |
| Class II obesity | 40 (5.5%) |
| Morbid obesity | 19 (2.6%) |
| **Fasting Plasma Glucose** |  |
| Normal | 573 (49.5%) |
| Prediabetes | 314 (27.1%) |
| Diabetes | 271 (23.4%) |
| **Hypertension** | 236 (20.4%) |
| Systolic BP | 127 (118–137) |
| Diastolic BP | 80 (72–85) |
| **ICU admission** | 104 (9%) |
| **Death** | 40 (3.5%) |

BMI: body mass index, BP: blood pressure, COVID-19: coronavirus disease 2019, ICU: intensive care unit.
<sup>a</sup>Values are expressed as medians (interquartile ranges) or as numbers (%).

### Characteristics of patients admitted to ICU

The characteristics of patients admitted to ICU were compared with those who did not require ICU care and are presented in **Table 2**. Among patients admitted to the ICU, the median (IQR) blood glucose level was 147 (108–208) mg/dL. In contrast, the remaining 1054 patients who did not require ICU care had significantly lower blood glucose levels (median [IQR]: 98.6 [86.2–118] mg/dL; *p*-value < 0.001). The patients admitted to the ICU were also significantly older (median age [IQR]: 54 [46.4–63.4] years) than those who did not require ICU care (median age [IQR]: 39.3 [30.7–50.4] years; *p*-value < 0.001). The median BMI of patients admitted to ICU was also statistically significantly higher than in patients who did not require ICU care (median [IQR]: 27.5 [25.3–31.4] kg/m^2^ vs 26 [23–29] kg/m^2^, respectively; *p*-value <0.001). There was also a significantly higher proportion of patients with hypertension among those admitted into ICU than those who were not (16% vs 8.1%: *p*-value = 0.004).

**Table 2:** Comparison of vitals and laboratory measurements between patients with COVID-19 with and without ICU admittance.

| Characteristic | ICU admittance<sup>a</sup> |  | *p*-value |
| --- | --- | --- | --- |
|  | No (*N* = 1054) | Yes (*N* = 104) |  |
| **Age, years** | 39.3 (30.7–50.4) | 54.0 (46.4–63.4) | <0.001* |
| **Sex – Male** | 274 (26%) | 66 (63.5%) |  |
| **Body Mass Index, kg/m<sup>2</sup>** | 26 (23–29) | 27.5 (25.3–31.4) | <0.001*** |
| Normal weight | 253 (95.1%) | 13 (4.9%) | 0.006** |
| Overweight | 270 (88.8%) | 34 (11.2%) | 0.006** |
| Class I obesity | 83 (84.7%) | 15 (15.3%) | 0.006** |
| Class II obesity | 35 (87.5%) | 5 (12.5%) | 0.006** |
| Morbid obesity | 15 (78.9%) | 4 (21.1%) | 0.006** |
| **Fasting Plasma Glucose, mg/dL** | 98.6 (86.2–118) | 147 (108–208) | <0.001*** |
| Normal | 554 (96.7%) | 19 (3.3%) | <0.001** |
| Prediabetes | 295 (93.9%) | 19 (6.1%) | <0.001** |
| Diabetes | 205 (75.6%) | 66 (24.4%) | <0.001** |
| **Hypertension** | 47 (8.14%) | 24 (16%) | 0.004*** |
| Systolic BP, mm Hg | 127 (118–136) | 126 (116–142) |  |
| Diastolic BP, mm Hg | 80 (72–85) | 73.5 (65.6–82) | <0.001** |

\*Student's *t* test.
\*\*Chi-squared test.
\*\*\*Mann-Whitney *U* test.
BP: blood pressure, COVID-19: coronavirus disease 2019, ICU: intensive care unit.
<sup>a</sup>Values are expressed as medians (interquartile ranges) or as numbers (%).

All classifications of BMI that were higher than normal were associated with admission to the ICU (*p*-value = 0.006). The association between obesity, diabetes and hypertension was tested with the chi-square test, which revealed a relationship between obesity and diabetes (*p*-value < 0.001); we therefore decided to construct two predictive models for ICU progression.

### Univariate analysis for ICU admission

Potential predictors for admission into ICU, including age, BMI and diabetes status, were analysed by univariate logistic regression (**Table 3**). Using BMI as the only predictor of ICU admission, the likelihood of admission was approximately 2.5 fold higher in patients who were overweight compared with patients with normal weight (odds ratio [OR] = 2.45; 95% CI: 1.2–64.74; *p*-value = 0.008). Patients with obesity were 3.5 and 5.2 times more likely to be admitted to ICU than patients with normal weight, depending on whether they had class I or morbid obesity, respectively (OR: 3.51 [95% CI: 1.60–7.69 for class I obesity; and OR: 5.18 [95% CI: 1.50–17.85] for morbid obesity). Diabetes was also an independent predictor for admission into the ICU (OR = 9.38; 95% CI: 5.49–16.02; *p*-value <0.001). Other variables, including age, sex, hypertension, prediabetes and class II obesity were not significant predictors for ICU admission.

**Table 3:**
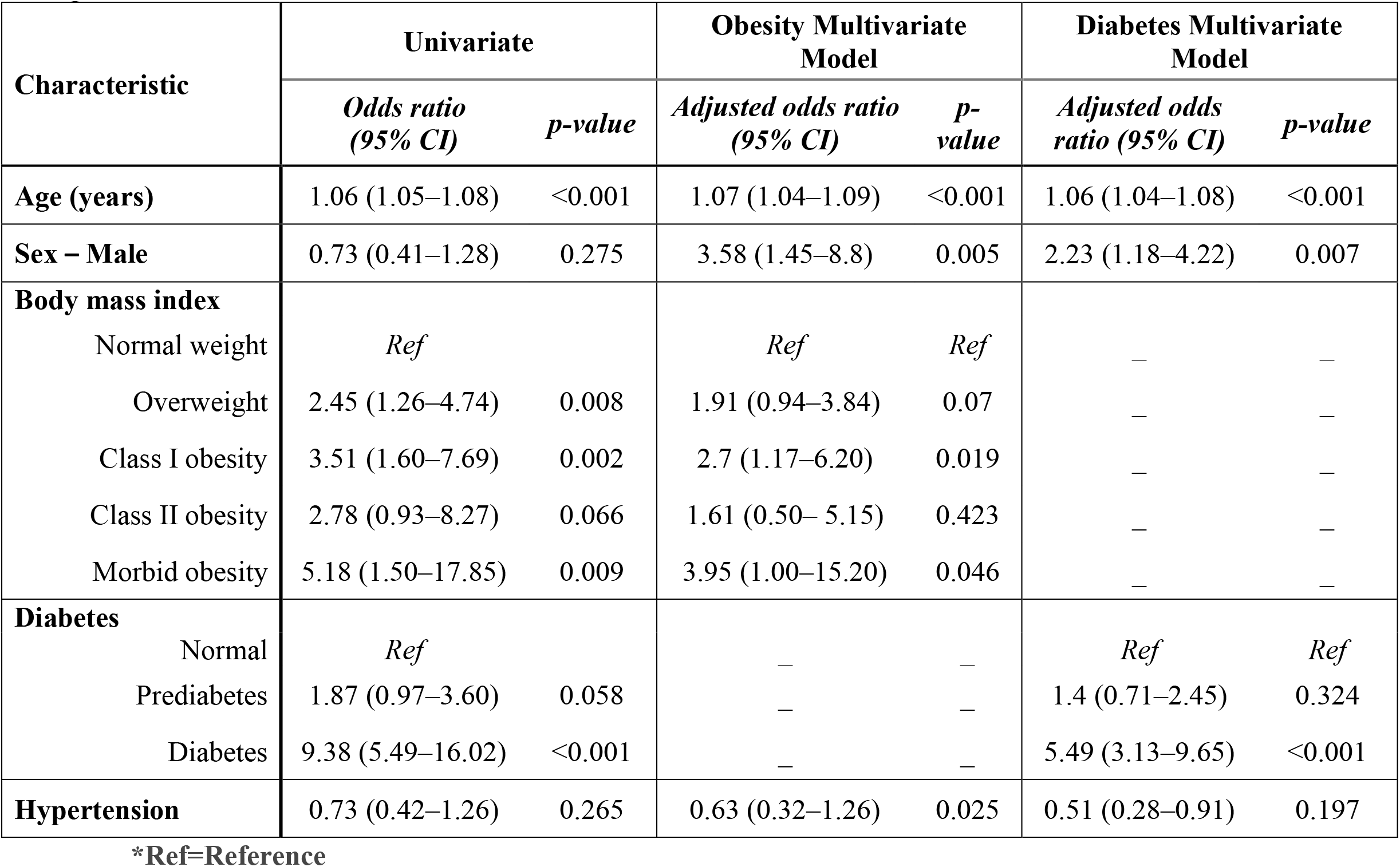
Univariable and multivariable analyses for obesity and other comorbidities, odds ratio adjusted by age and gender.

### Multivariate analysis for ICU admission

Considering the association between obesity and diabetes and to exclude the potential confounding effect of these two variables on the outcome, we constructed two multivariate logistic regression models of prediction to more accurately assess the associations of each of these two conditions with ICU admittance (**Figure 1**). In the first multivariate analysis model, where obesity was assessed, BMI (adjusted for age and sex) significantly associated with admittance to the ICU. In patients who were overweight, had class I obesity and morbid obesity, the adjusted OR (AOR) were 1.91 (95% CI: 0.94–3.84), 2.7 (95% CI: 1.17–6.20), and 3.95 (95% CI: 1.00–15.2), respectively (**Figure 1**). Sex was also identified as a risk factor for ICU admission (AOR: 3.58 [95% CI: 1.45–8.8]). On the other hand, hypertension reduced the risk of ICU admission by approximately 40% (AOR: 0.63; 95% CI: 0.32–1.26; *p*-value = 0.025).

In the second multivariate analysis model of prediction, diabetes (adjusted for age and sex) was associated with an increased the risk of ICU admittance (AOR = 5.49; 95% CI: 3.13–9.25, p-value ≤ 0.001); however, patients with prediabetes did not have an increased risk of being admitted to ICU (AOR: 1.4 [95% CI: 0.71–2.45]; **Figure 1**). Both multivariate models were validated by calculating the area under the receiver operating characteristic (ROC) curve, which were above 0.800 for both multivariable models, indicating a good model discrimination **(Figure 2 and 3)**.

**Figure 1:**
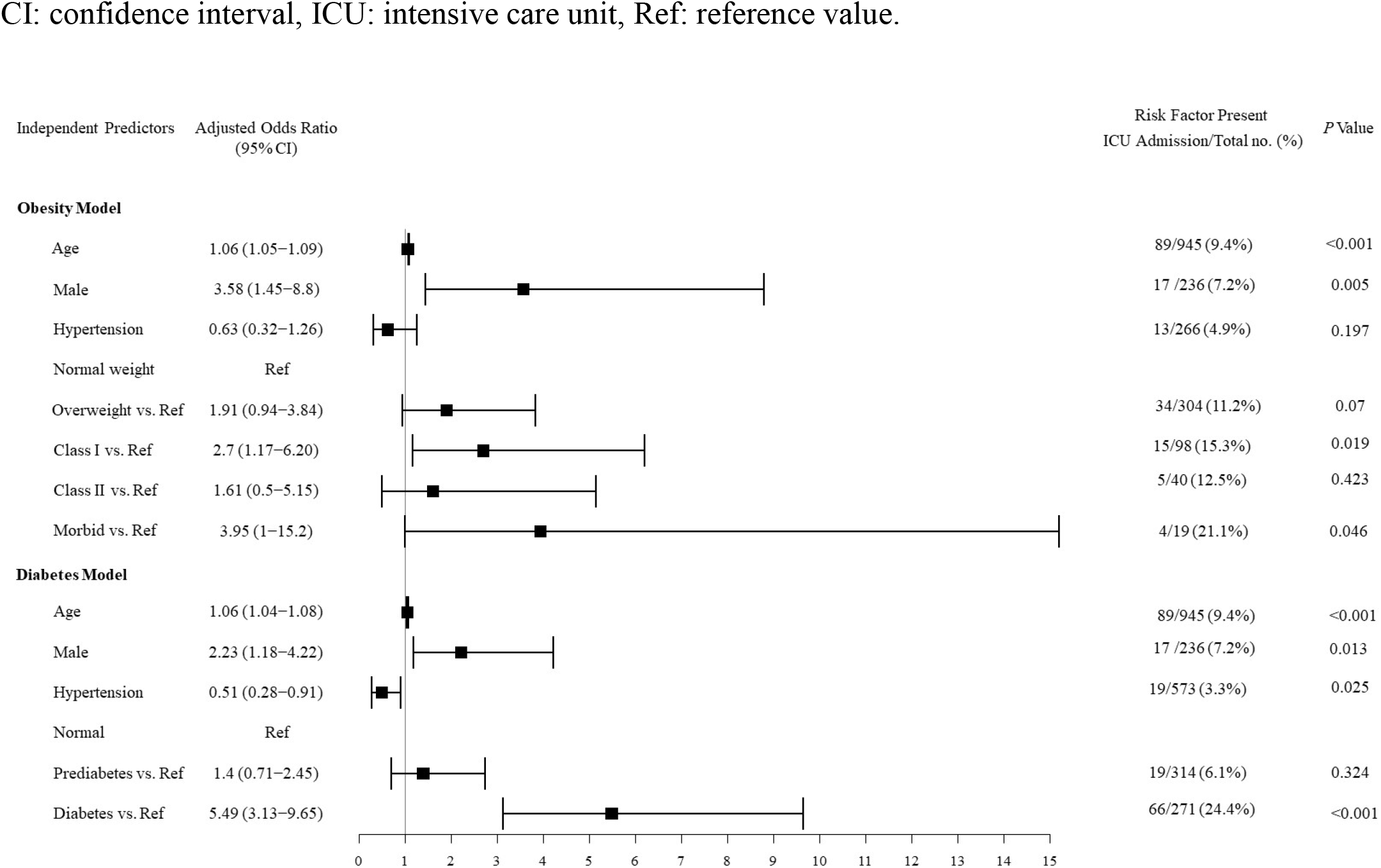
Independent predictors of ICU admission from multivariate logistic regression analysis. CI: confidence interval, ICU: intensive care unit, Ref: reference value.

**Figure 2:**
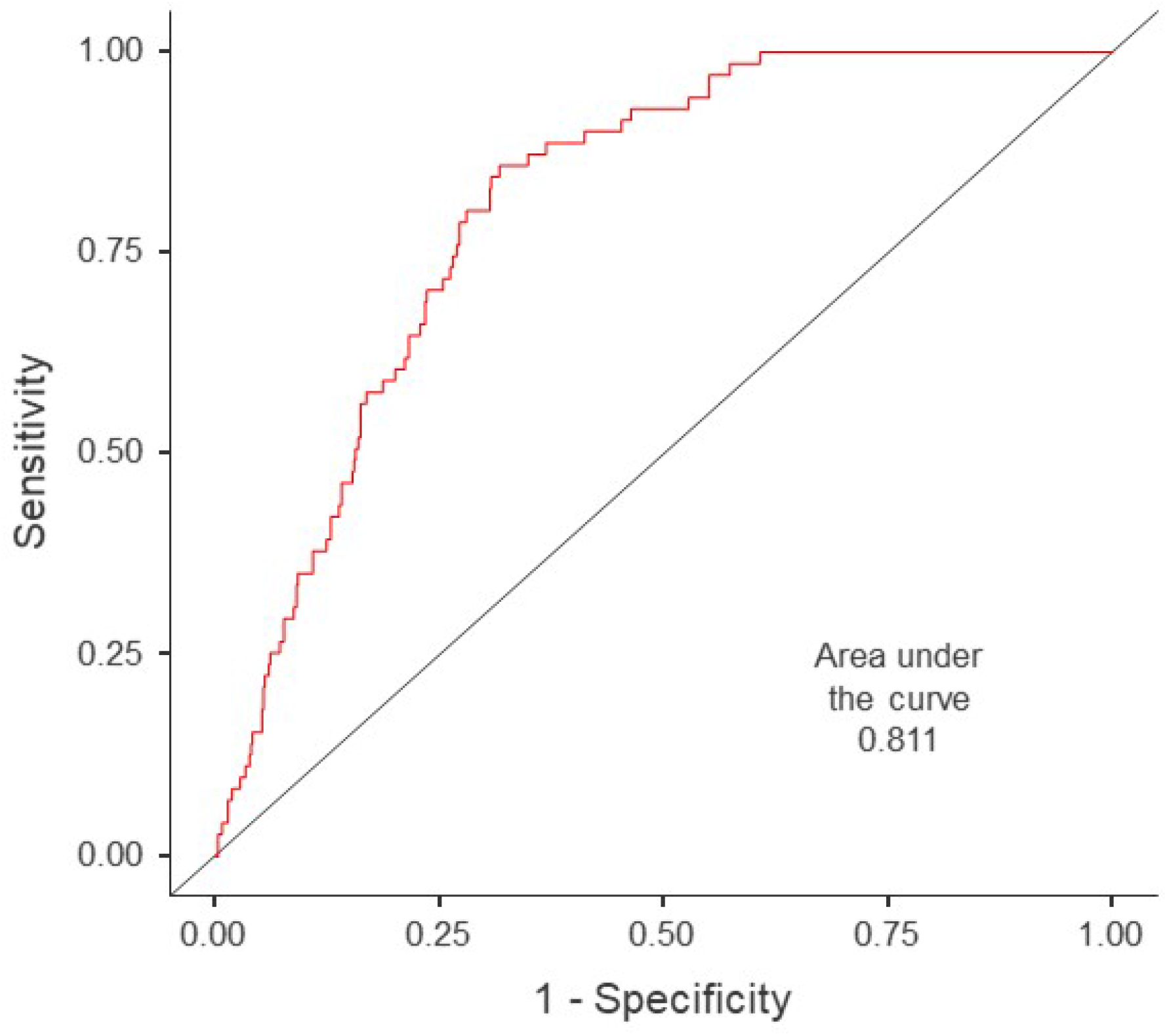
Receiver operating characteristic curve for the BMI multivariate model of prediction.

**Figure 3:**
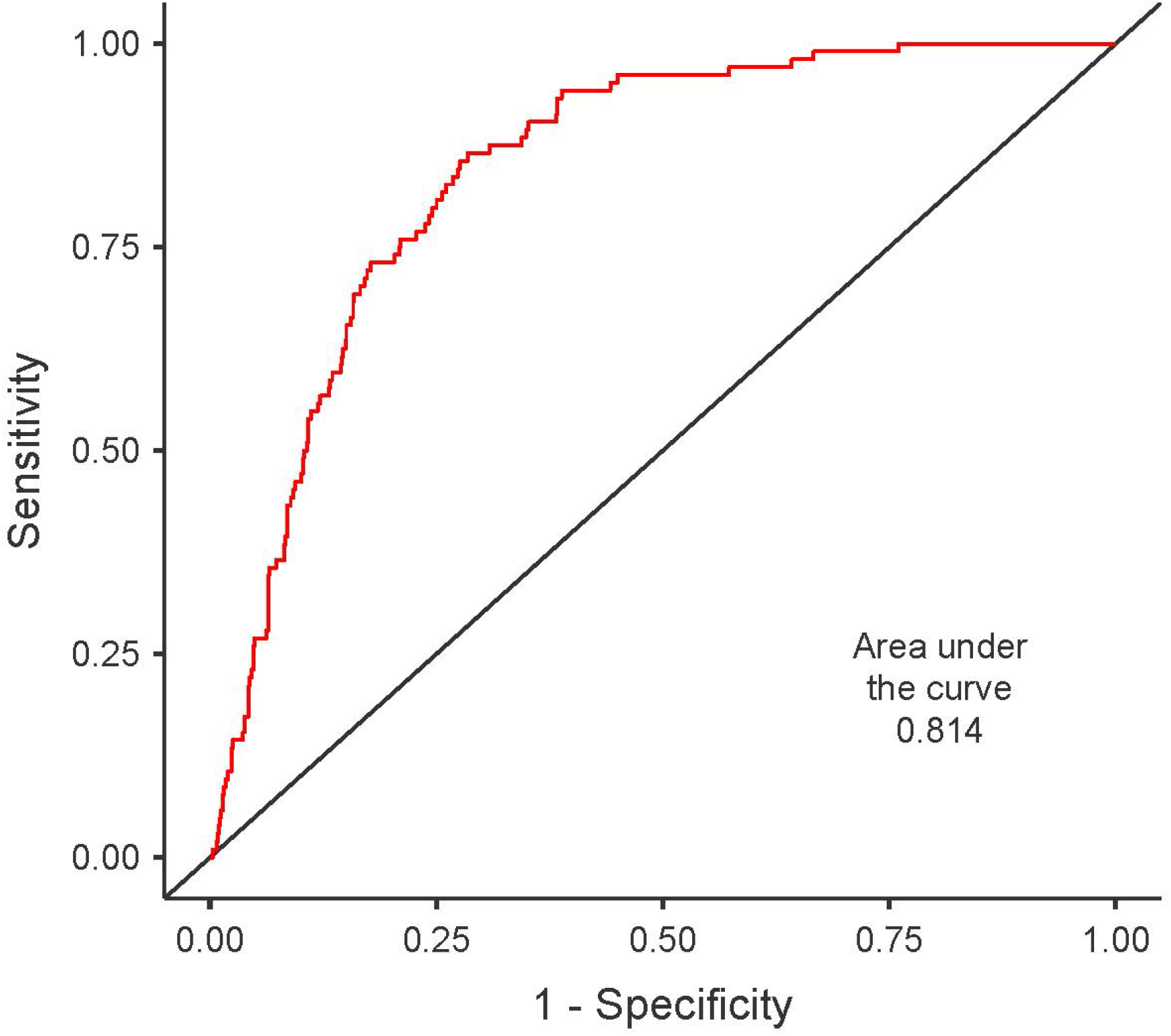
Receiver operating characteristic curve for the diabetes multivariate model of prediction.

## Discussion

This was the first retrospective cohort study in Kuwait and the Middle East which analysed the potential association of obesity and diabetes with COVID-19 disease severity. All patients in our cohort were admitted to a single centre, where they underwent standardised sets of investigations and received standardised treatment protocols.

In this study, univariate logistic regression analysis revealed that BMI and diabetes were independent factors associated with admission to the ICU. After adjusting for confounding factors, further multivariate analysis confirmed these results, showing that in our population, the presence of diabetes and higher BMI were associated with ICU admittance. In concordance with our findings, a study from a New York Hospital by Lighter et al. demonstrated that patients with COVID-19 who had a BMI between 30 and 34.9 were two times more likely to be admitted to the acute critical care unit than non-obese patients (4). In the same vein, in a retrospective cohort of 124 patients in France, Simonnet et al. found that obesity prevalence was high among patients admitted to the ICU (5). Although obesity is not as prevalent in China as in the Middle East, Peng et al. obtained similar results in a retrospective analysis of 112 patients with COVID-19, who were admitted to Wuhan Union Hospital in Wuhan, China (6). When patients were divided into two groups according to the severity of the disease (critical and general), researchers observed that the BMI of the critical group was significantly higher than that of the general group (25.5 [23.0–27.5] kg/m^2^ vs 22.0 [20.0–24.0] kg/m^2^, respectively; *p*-value = 0.003) (6).

In our study sample, class II obesity was not significantly associated with admittance to the ICU, though this could be attributed to the small sample size and the fact that a large proportion of patients in this class had no symptoms.

The association between diabetes and admission to the ICU seen in our cohort by multivariate analysis of prediction was not entirely surprising. A greater risk of severe COVID-19 in patients with diabetes has been reported in several studies; the higher risk of respiratory infections has been attributed to the compromised immune system, especially the innate immunity, of patients with diabetes. Even transient hyperglycaemia may temporarily affect the innate immune response to infection (7).

The exact mechanism through which obesity and diabetes contribute to severe outcomes among patients with COVID-19 is not yet known. However, one explanation could be related to the fact that expression of angiotensin-converting enzyme 2, the functional receptor for SARS-CoV, is up-regulated in patients with obesity and diabetes (8). Furthermore, obesity and diabetes are believed to be linked with dysregulated lipid synthesis and clearance, which can initiate or aggravate pulmonary inflammation and injury (9). Further mechanistic studies on this matter are needed to elucidate the mechanism by which diabetes and obesity contribute to disease severity and poor outcomes among patients with COVID-19.

The novel COVID-19 pandemic has created an unprecedented challenge in health care, exacerbating the unavailability of medical resources throughout the world. Bariatric surgeons have witnessed the rise of obesity and diabetes in epidemic proportion and have come to realise the effect of this rise on the medical care system. In our study, diabetes and BMI were associated with more severe COVID-19 outcomes, assessed as ICU admittance of hospitalized patients. We acknowledge that our study had some limitations. Given its retrospective nature, the unavailability of data as a result of omission or inadequate recording was a major limitation of this study. Another limitation of this study was the relatively higher ratio of male to female patients, which may limit the generalisability of the results to the population. The relatively small size of our sample population was another limitation. Thus, larger, multicentre studies are needed to confirm our findings and provide more robust scientific evidence. Nevertheless, our findings indicate that more patients with obesity and diabetes are likely to be admitted to the ICU as the pandemic continues. Hence, patients with COVID-19 with underlying obesity or diabetes must be categorised as a highrisk group.

## Data Availability

All data included in the manuscript can be made available, on request

## Acknowledgements

We would like to thank Dr. Diana Marouco, PhD, for the editorial support and Bruno Siegel Guerra for the statistical provided.

## Competing Interests

All authors declare no competing interests of any form.

## Notes

### Competing Interest Statement

The authors have declared no competing interest.

### Funding Statement

Funding for this project was provided by Kuwait Foundation for the Advancement of Science

### Author Declarations

Ethical approval was obtained from the Ministry of Health Kuwait

